# Prevalence and Functional Outcomes of Post-Exertional Malaise among Adults with prior COVID-19: Results from a Representative Survey of New York City Residents

**DOI:** 10.64898/2026.08.31.26356614

**Authors:** Samuel E. Packard, Therese Russo, Jannae Parrott, Julia Sisti, Amanda Lans

**Author notes:** Corresponding author: Samuel E. Packard.

## Abstract

**Objective:** To estimate the prevalence of Post-Exertional Malaise (PEM) among adults with prior COVID-19 and associated mental health and disability outcomes.

**Methods:** We conducted a cross-sectional analysis of data from a survey of 9,620 adults with prior COVID-19 in New York City, collected May – June 2024. PEM was measured with the DePaul Symptom Questionnaire – Post Exertional Malaise, categorized by symptom duration (< 14 vs. ≥14 hours). Weighted prevalence estimates were stratified by socio-demographic and clinical characteristics. Modified Poisson regression was used to assess the association of PEM with depression, anxiety, and disability.

**Results:** The prevalence of PEM symptoms was 20.9% overall and 4.0% with symptom duration ≥ 14 hours, representing over 800,000 New Yorkers affected and over 150,000 who meet a diagnostic criterion for ME/CFS. PEM prevalence was higher among women, transgender and non-binary adults, people of color, and lower educational attainment, chronic comorbidities, or disabilities. PEM was associated with 3 - 4 times higher prevalence of mental health outcomes and 4 - 5 times higher disability scores.

**Conclusions:** PEM symptoms were common and strongly associated with disability and adverse mental health. Screening, pathways to care, and supportive policies are needed to mitigate long-term consequences, particularly among marginalized populations.

## 1. Introduction

Many people do not fully recover from SARS-CoV-2 infection within 3 months, remaining below their pre-infection baseline health for months or years after acute infection^1–3^. In New York City (NYC), one in four adults infected with COVID-19 between 2020 – 2023 reported related long-term physical or mental health issues^4^. While this represents a broad range of sequelae, a survey of post-acute symptoms in this population found that fatigue and decreased exercise tolerance were the most commonly reported^5^. The prevalence of these consequential yet non-specific symptoms indicates a need to better characterize the conditions they may represent.

Post-exertional malaise (PEM), also known as post-exertional neuroimmune exhaustion or post-exertional symptom exacerbation, is one such symptom often mischaracterized as general fatigue in clinical care and research^6^. PEM is an exacerbation of symptoms, including but not limited to extreme fatigue, cognitive dysfunction, pain, muscle weakness, and influenza-like illness, that occurs after physical or cognitive exertion. Unlike a typical response to physical or cognitive effort in otherwise healthy individuals, the symptom exacerbation characteristic of PEM (often described as a “crash”) can occur in response to even mild exertion, can be delayed by up to 12 to 48 hours, and may last for days, weeks, or longer, resulting in significant functional impairment^7,8^. The hallmark symptom of myalgic encephalomyelitis/chronic fatigue syndrome (ME/CFS), a chronic, disabling neurological disease often preceded by bacterial or viral infection^6,7^, PEM has also been reported among people experiencing long-term illness after acute COVID-19 (long COVID), indicating that the SARS-CoV-2 virus may be among the etiologic agents that cause ME/CFS or may trigger a distinct illness that presents with ME/CFS-like symptoms^9–11^. Given its disabling effects, estimating the prevalence of PEM and associated burden of illness is critical to mitigating the sustained public health impacts of the pandemic ^12,13^.

Despite the implications for public health, PEM is rarely identified in common epidemiologic data sources such as routine disease reports, electronic health records, laboratory data, or syndromic surveillance. Diagnosis of PEM requires clinical evaluation alongside assessment of patient-reported information, which is not practical for public health surveillance. The DePaul Symptom Questionnaire – Post Exertional Malaise short form (DSQ-PEM) is a screening tool which measures the frequency and severity of five symptoms selected from the 54-item DePaul Symptom Questionnaire due to their high sensitivity and specificity for identifying PEM and has been recommended by patient advocates and researchers for population health applications^14–18^. Prior studies using the DSQ-PEM have suggested that PEM symptoms may present in 20 - 25% of COVID-19 cases after 3-6 months, and half of all people who have long COVID^19–22^.

In this study, we estimate the prevalence of PEM among adult New York City residents with prior COVID-19 using the DSQ-PEM in a large, population-representative survey. First, we calculate survey-weighted estimates of the number and proportion of adult New Yorkers with prior COVID-19 who met criteria for PEM symptoms and those who experienced prolonged (≥ 14 hour) symptom duration. We then calculate stratified estimates by socio-demographic characteristics and assess the association of PEM symptoms with mental health outcomes and disability, adjusting for potential confounders.

## 2. Methods

### 2.1. Study Design and Population

We used data from the baseline survey of the Long-term Outcomes of New Yorkers with COVID-19 Study, a cohort study conducted by the New York City Department of Health and Mental Hygiene (NYC Health Department) consisting of four surveys fielded over two years to assess the long-term outcomes of SARS-CoV-2 infection among adult NYC residents. From the May - June 2024, panelists in the NYC Health Panel^23^ (n = 34,811) were invited to an eligibility screener for the baseline survey; criteria included residence in NYC, age 18 or older, and prior COVID-19 infection. Prior infection was assessed by the question “Do you think you have ever had COVID-19?” with five affirmative response options: Positive rapid test, positive PCR test, positive antibody test, healthcare provider diagnosis, or “some other reason”. The survey was available to complete by computer, mobile device, or computer-assisted telephone interview in English, Spanish, Russian, or Chinese.

All panelists who met the above criteria, completed the baseline survey, and had complete data for exposure and outcome variables (PEM symptoms, probable depression, probable anxiety, and disability) were included in the present study. Estimates were weighted to the demographics of the target population of adult New Yorkers with prior COVID-19 per the NYC Health Department’s 2023 Community Health Survey using borough, age group, sex at birth, race/ethnicity, educational attainment, and presence of children in the household.

### 2.2. Measures

The DSQ-PEM ascertains self-reported frequency and severity of five symptoms experienced after exertion over the past six months: Dead, heavy feeling after starting to exercise; Next day soreness or fatigue after non-strenuous, everyday activities; Mentally tired after the slightest effort; Minimum exercise makes you physically tired; Physically drained or sick after mild activity. PEM was defined as reporting at least one of the five symptoms at both a severity of moderate or greater and a frequency of about half the time or more^18^. Symptom duration following a triggering activity was dichotomized as mild/moderate (<14 hours) and prolonged (≥14 hours). Prolonged symptom duration is relevant as a diagnostic criterion of ME/CFS^18^.

Probable depression and anxiety were defined as a score of ≥ 3 on the Patient Health Questionnaire 2 (PHQ-2) and Generalized Anxiety Disorder 2 (GAD-2), respectively^24,25^. Disability was measured using the 12-item version of the World Health Organization Disability Assessment Schedule 2.0 (WHODAS-2). The global WHODAS-2 score ranges from 0 (no disability) to 48 (full disability), with domain scores ranging from 0 - 8 for each of 6 subdomains, calculated using simple scoring with mean substitution for missing items^26^. Long COVID was defined as ever experiencing new or exacerbated symptoms lasting 3 months or longer after acute COVID-19.

Self-reported socio-demographic characteristics included age, gender identity, race, ethnicity, and educational attainment. Neighborhood poverty was defined by the proportion of households in the respondent’s ZIP code below the federal poverty level according to the 2016 – 2020 American Community Survey. Self-reported indicators of health history included number of COVID-19 infections, year of first infection, number of COVID-19 vaccines, number of pre-existing chronic conditions (self-reported diagnosis of diabetes, hypertension, chronic lung disease, cardiovascular disease, cancer, chronic kidney disease, chronic liver disease, neurological disease, mood disorder, or immunodeficiency) and pre-existing disability (difficulty with hearing, vision, cognition, mobility, self-care, and/or independent living).

### 2.3. Statistical analysis

First, we described the characteristics of the study sample with unweighted counts and weighted percentages. Then, we estimated the weighted prevalence of PEM symptoms (overall and with prolonged duration) by selected socio-demographic and health characteristics with Chi-squared tests to assess differences. Weighted counts were estimated to the nearest 1,000. We then fit a series of regression models to assess the association between a 3-level categorical variable for PEM (No PEM, PEM with mild/moderate duration, and PEM with prolonged duration) and three outcomes: probable depression, probable anxiety, and disability. Probable depression and probable anxiety were modeled separately as binary outcomes in survey-weighted generalized linear models with Poisson distribution and log link function with robust standard errors to estimate relative risks (RR)^27^. For disability, we fit survey-weighted linear regression models with WHODAS-2 score as a continuous outcome variable, estimated marginal predicted mean values by PEM symptom category, and reported the ratio of mean WHODAS-2 scores for PEM symptoms (mild/moderate duration and prolonged duration) compared to the referent group of no PEM symptoms. For each model, an unadjusted model was compared to a model adjusted for age group, gender, race/ethnicity, educational attainment, neighborhood poverty level, number of prior COVID-19 vaccines, year of first COVID-19 infection and number of prior COVID-19 infections, number of pre-existing chronic conditions, and pre-existing disability as covariates. Missing data were handled using the Missing Covariate Indicator Method (MCIM), as all covariates had < 5% missingness and MCIM introduces minimal bias under such conditions^28^.

We conducted two sensitivity analyses. To assess the association of symptom duration with mental health and disability among people with PEM, we reproduced the models excluding participants with no PEM, using mild/moderate duration as the referent group. To address potential confounding by pre-existing conditions, we excluded participants with a history of diagnosed mood disorders from models of mental health outcomes and excluded participants with pre-existing disability from models of disability outcomes.

Descriptive analyses were performed in SAS 9.4 (SAS Institute Inc., Cary, NC, US). Statistical models were programmed in R 4.3.2 using the survey and marginaleffects packages^29–30^. This study was approved by the NYC Department of Health Institutional Review Board (Protocol #24 – 007).

## 3. Results

### 3.1. Participant Characteristics

Participant characteristics are shown in Table 1. A total of 9,620 participants met inclusion criteria for the study, representing 4.2 million adult New York City residents with at least one COVID-19 infection prior to June 2024. A total of 220 survey responses (2.2%) were excluded due to missing values for PEM, mental health, or disability. Overall, 90% of respondents had clinical or laboratory evidence of prior COVID-19; 43% reported a positive PCR test, 40% reported a positive rapid test, 7% reported a positive antigen test or physician diagnosis in absence of PCR or rapid test, and 10% reported some other reason (Supplementary Table S-1).

**Table 1:**
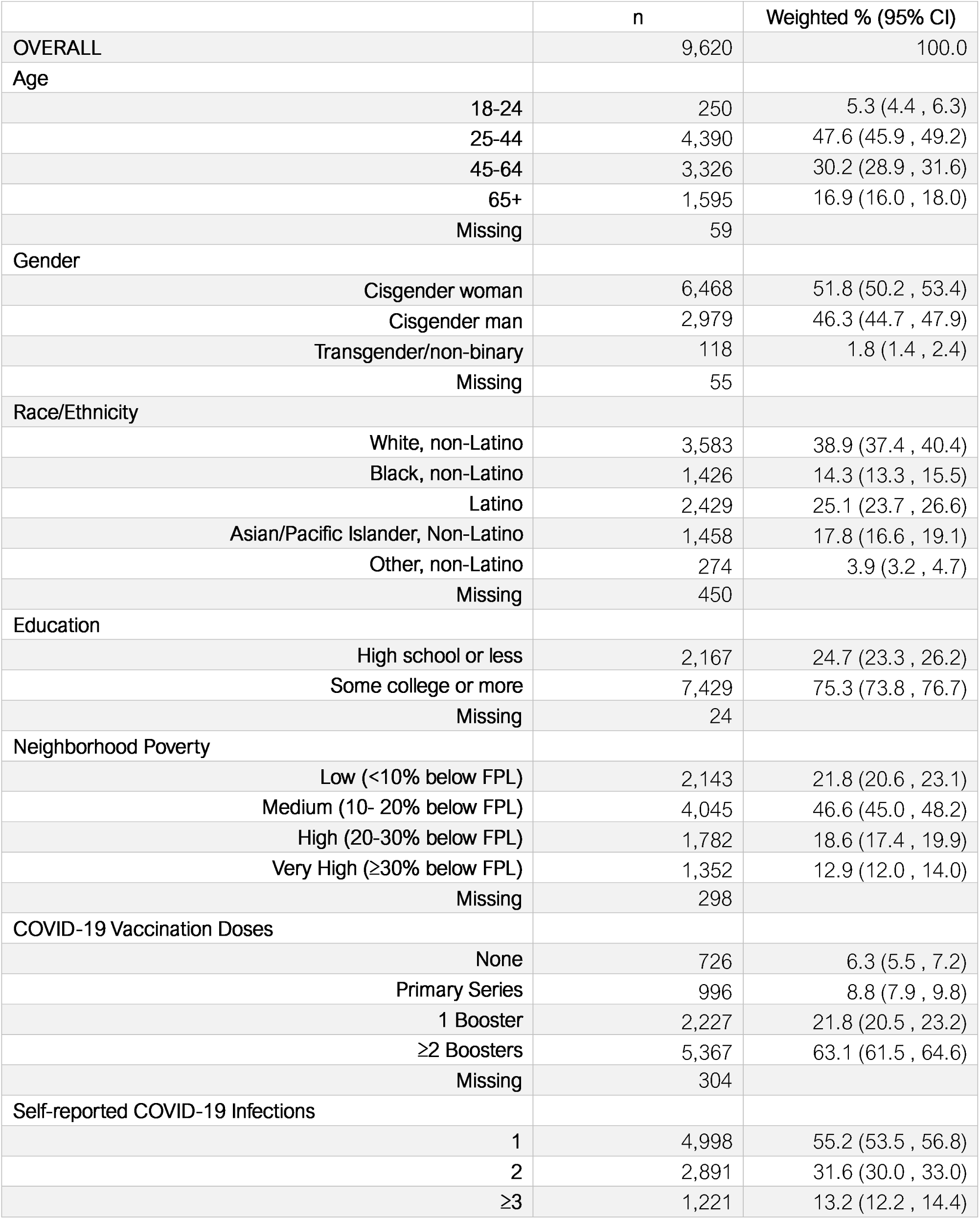
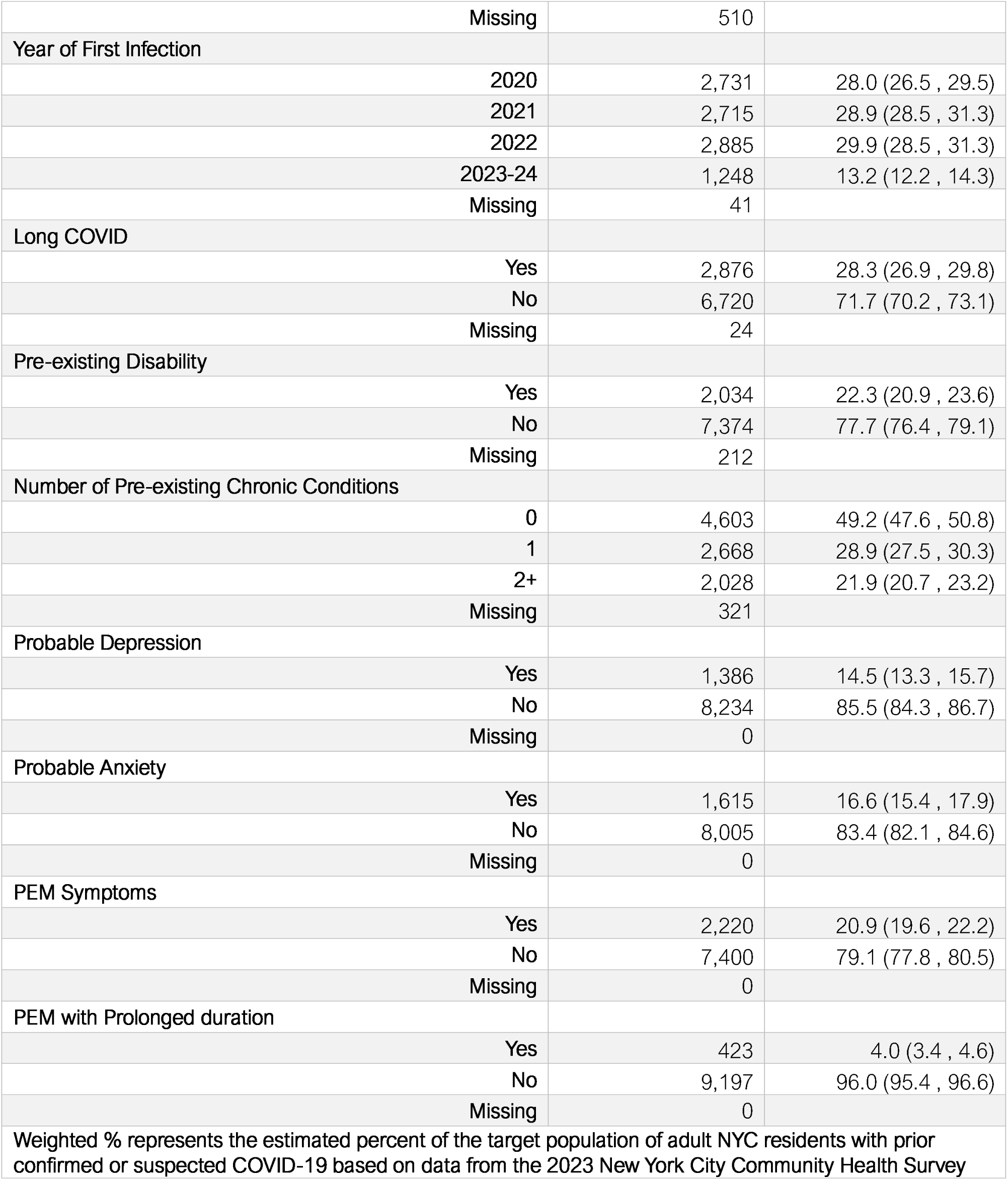
Selected characteristics of the study sample: New York City, May – June 2024.

### 3.2. Prevalence of post-exertional malaise

Prevalence of PEM symptoms by socio-demographic and health characteristics are shown in Table 2. Among adults with prior COVID-19, the prevalence of PEM symptoms was 20.9% and PEM symptoms with prolonged duration was 4.0%, representing approximately 841,000 and 160,000 New Yorkers, respectively. Among adults with a history of long COVID, the prevalence of PEM was 42.3%. The prevalence of PEM symptoms was higher among cisgender women and transgender adults (compared to cisgender men), Black, Latino, and Other race (compared to White, non-Latino adults), people with a high school education or less, people living in higher poverty neighborhoods, people infected earlier in the pandemic, and people with multiple COVID-19 infections, fewer vaccine doses, pre-existing disability, pre-existing chronic conditions, and long COVID. Statistically significant differences were not found by age group or vaccination status. While fewer statistically significant between-group differences were found in the prevalence of PEM symptoms with prolonged duration, it was observed to be more prevalent among people with long COVID, multiple COVID-19 infections, infections earlier in the pandemic, pre-existing chronic conditions, and pre-existing disability, and to differ significantly by age group and race/ethnicity.

**Table 2:**
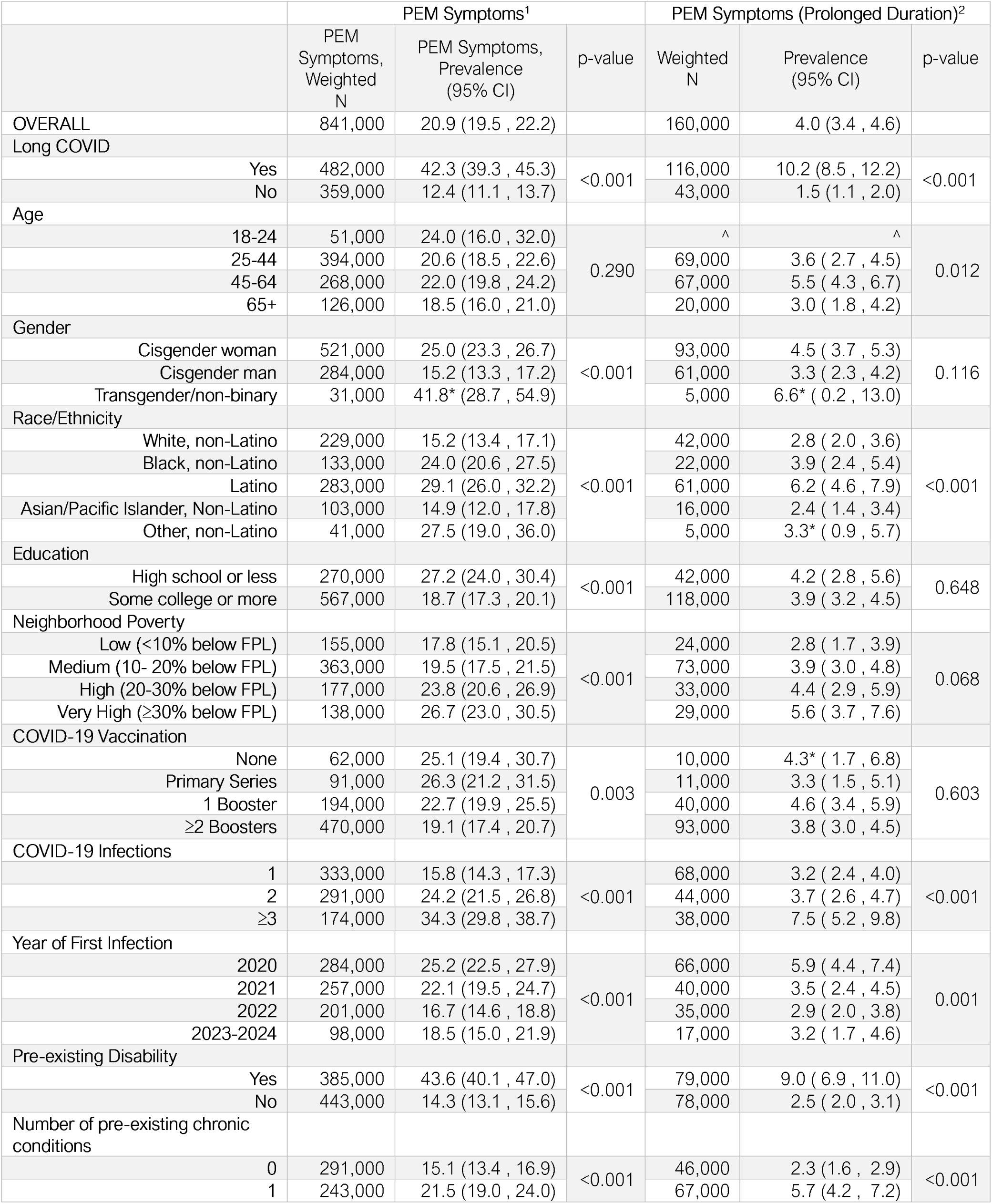
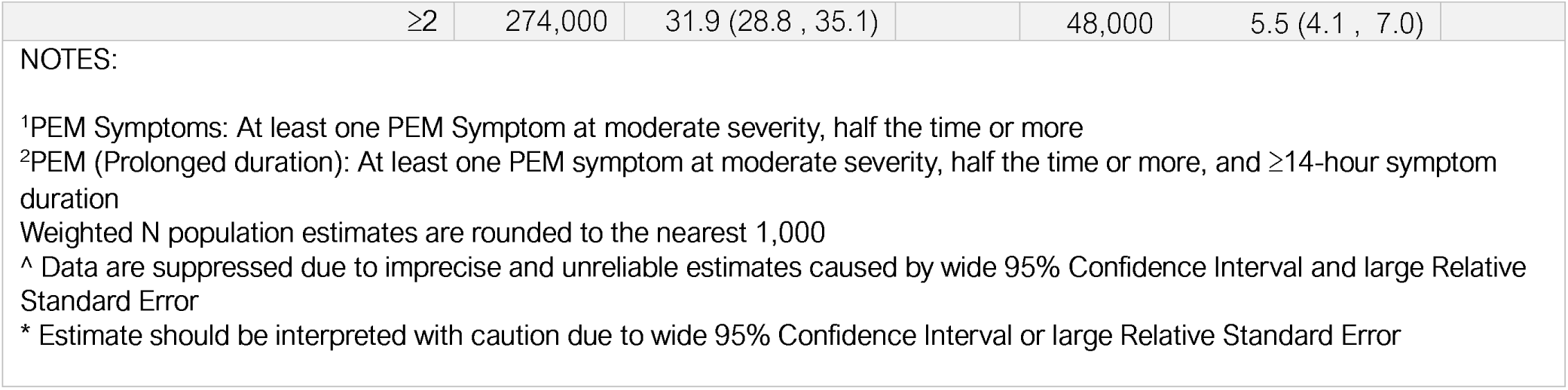
Prevalence of Post-Exertional Malaise Symptoms by Selected Characteristics among adult New York City residents with prior COVID-19, May – June 2024.

### 3.3. Association of post-exertional malaise with mental health and disability

The overall prevalence of probable depression and anxiety in the study sample was 14.5% (95% CI: 13.3,15.7) and 16.6% (95% CI: 15.4,17.9), respectively (Table 1).Compared to people without PEM symptoms, PEM was associated with 3-4 times the risk of these outcomes after adjusting for potential confounders (Table 3). Unadjusted mean WHODAS-2 global scores were 3.1 (95% CI: 2.9,3.4) among people reporting no PEM symptoms, 14.2 (95% CI: 13.4,15.0) for PEM symptoms with mild/moderate duration, and 19.1 (95% CI: 17.3,21.0) for PEM symptoms with prolonged duration (Supplementary Table S-2). WHODAS-2 global scores were on average 3.4 (95% CI: 3.1,3.7) times higher and 4.7 (95% CI: 4.1,5.3) times higher among people with PEM symptoms with mild/moderate and prolonged duration, respectively (Figure 1). PEM symptoms were also associated with higher subdomain scores across all 6 domains of disability.

**Figure 1:**
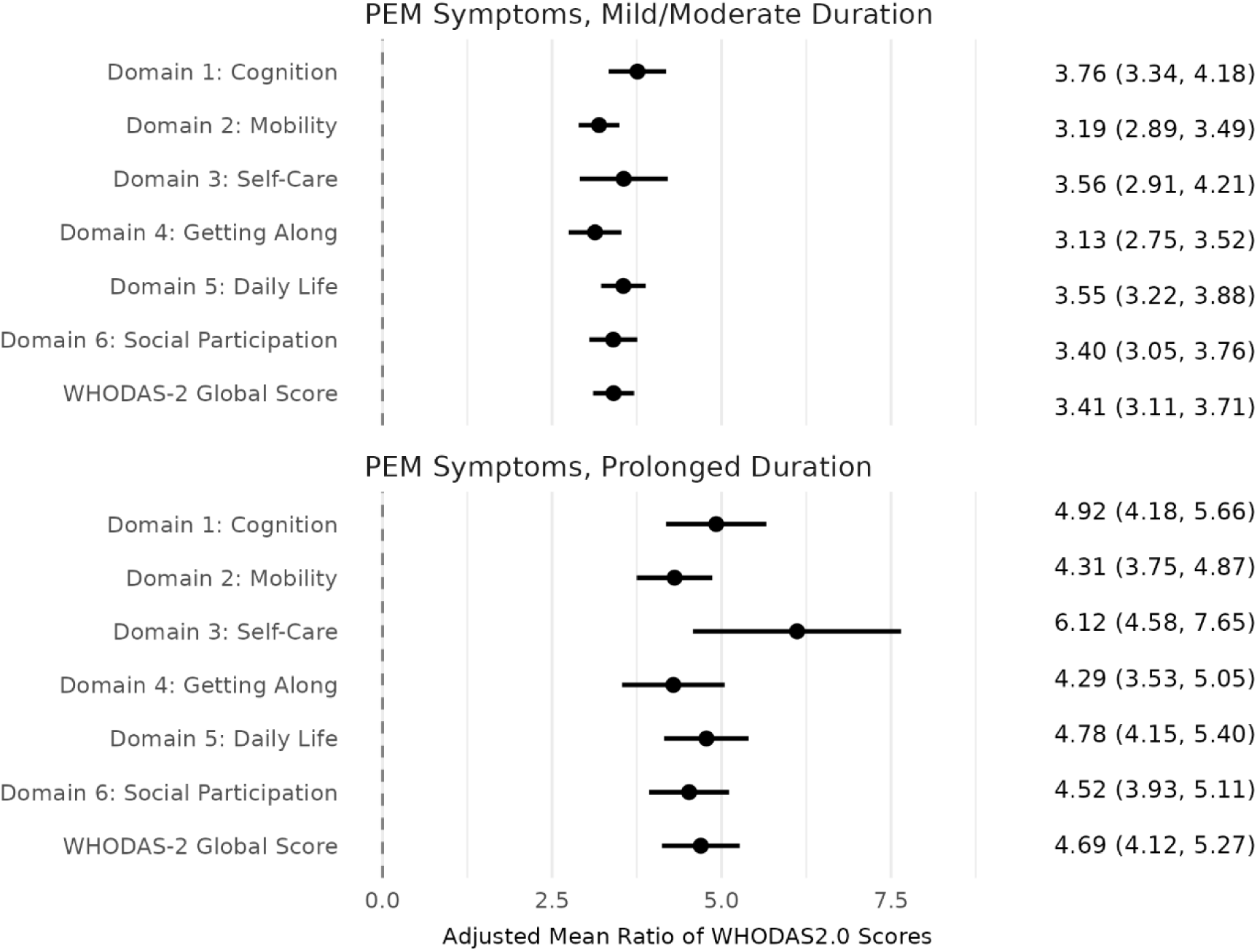
Association of post-exertional malaise symptoms with disability among adult New York City residents with prior COVID-19, May – June 2024. Notes: Estimates are shown as the Adjusted Mean Ratio of WHODAS-2 scores and 95% Confidence Intervals. The referent group for each estimate is No PEM Symptoms. Estimates are adjusted for age group, gender, race/ethnicity, educational attainment, neighborhood poverty, number of prior COVID-19 infections, number of prior COVID-19 vaccines, year of first COVID-19 infection, pre-existing chronic condition, and pre-existing disability.

**Table 3:** Association of post-exertional malaise symptoms with probable depression and anxiety among adult New York City residents with prior COVID-19, May – June 2024.

|  |  | Probable Depression | Probable Anxiety |
| --- | --- | --- | --- |
|  |  | aRR (95% CI) | aRR (95% CI) |
| A | No PEM | ref | ref |
|  | PEM (Mild/Moderate Duration) | 5.23 (4.41 – 6.21) | 4.32 (3.72 – 5.03) |
|  | PEM (Prolonged duration) | 6.28 (5.06 – 7.79) | 5.48 (4.53 – 6.62) |
| B | No PEM | ref | ref |
|  | PEM (Mild/Moderate Duration) | 3.46 (2.87 – 4.19) | 2.93 (2.47 – 3.46) |
|  | PEM (Prolonged duration) | 4.14 (3.23 – 5.29) | 3.67 (2.94 – 4.57) |
| Notes: Row A displays results from unadjusted models, while Row B displays results from models adjusted for age group, gender, race/ethnicity, educational attainment, neighborhood poverty, number of COVID-19 infections, year of first infection, COVID-19 vaccination history, pre-existing chronic conditions, and pre-existing disability. Probable depression was defined as a PHQ-2 score $\geq 3$. Probable Anxiety was defined as a GAD-2 score $\geq 3$. All models were fit using modified Poisson regression for binary outcome data with robust standard errors and weighted to the target population of adult NYC residents with prior confirmed or suspected COVID-19. |  |  |  |

### 3.4. Sensitivity Analyses

Association of symptom duration with study outcomes among adults with PEM symptoms are shown in Supplemental Tables S-3 and S-4. Compared to people reporting mild/moderate symptom duration, prolonged symptom duration was not associated with risk of probable depression, but risk of probable anxiety was 27% higher (95% CI: 1.07, 1.51) and mean WHODAS-2 global scores were 33% higher (95% CI: 1.18, 1.47). Statistical models excluding participants with a history of mood disorders demonstrated similar associations with mental health outcomes as the main analysis, while models excluding participants with a history of disability demonstrated notably stronger association with disability (Supplemental Table S-5 and Figure S-1).

## 4. Discussion

Nearly one million New Yorkers with prior COVID-19 may meet screening criteria for post-exertional malaise on the DSQ-PEM, representing one in five adults with prior COVID-19 and nearly half of adults with a history of long COVID. PEM symptoms were more prevalent among groups disproportionately affected by social and structural determinants of health, including Black and Latino New Yorkers, people of lower socio-economic status, cisgender women and transgender or gender non-binary adults, and people with pre-existing disabilities and chronic health conditions, highlighting significant health inequities. Furthermore, an estimated 4% of adults with prior COVID-19 reported PEM symptoms lasting 14 or more hours after a triggering activity, representing more than 150,000 New Yorkers who meet a major diagnostic criterion for ME/CFS. The prevalence of adverse functional outcomes was substantially higher in people reporting PEM symptoms, with stronger effect sizes among those who reported prolonged symptom duration following a triggering event. These findings demonstrate a substantial burden of PEM symptoms with implications for daily functioning and mental health that may persist long after an acute COVID-19 infection has resolved, underscoring a public health issue that warrants attention.

Our estimate of the overall prevalence of PEM (20.9%) is consistent with a multi-center U.S. cohort reporting prevalence of 24% ≥ 6 months after SARS-CoV-2 infection, a population-based study of adults from Germany reporting prevalence of 20.5% two years after infection, and an observational study of adults from the Netherlands reporting prevalence of 23.2% in women and 17.8% in men ≥ 3 months after infection^19–21^. Our estimate of the prevalence of PEM among people with a history of long COVID (42%) is consistent with a meta-analysis of 12 studies which found a pooled prevalence of 55% (95% CI, 38% - 71%) in post-acute COVID-19 syndrome^22^. Our prevalence estimate of PEM with prolonged duration (4.0%), a diagnostic criterion for ME/CFS, was similar to the 4.5% prevalence estimates of ME/CFS-like illness in people with prior SARS-CoV-2 infection reported by two recent multi-center studies^20,21^. While the instruments used to measure depression, anxiety, and disability in this study have not been used widely in research on PEM or ME/CFS, the elevated prevalence of these functional outcomes associated with PEM in our study are consistent with research demonstrating worse physical, mental, and quality of life outcomes among people experiencing PEM^31–32^.

These results have several strong public health implications. First, the prevalence of symptoms that may warrant clinical evaluation for PEM or ME/CFS underscores the need to promote monitoring, detection, and clear pathways for diagnosis and referral to care for these conditions. Second, the elevated risk for disability and adverse mental health supports the utility of the DSQ-PEM in identifying people who experience disruptive physical and cognitive fatigue in response to mild exertion, and may benefit from clinical support and symptom management even in the absence of a clinical diagnosis. In other words, PEM symptoms alone merit attention as a public health concern not limited to a specific diagnosable condition or etiology. Third, since PEM can significantly limit a person’s ability to fulfill important life roles, including work, education, and caregiving, inclusive and supportive policies are needed to offset potential impacts to social and economic well-being such as workplace accommodations, access to disability evaluations, and referral to public benefits and social services. Fourth, there is an urgent need to increase awareness of PEM among the general public and improve training for health providers. Limited awareness contributes to stigma, while insufficient clinical training can result in patients with PEM being undiagnosed, misdiagnosed, and prescribed harmful treatments. In particular, the strong association of PEM with symptoms of depression and anxiety presents a danger of PEM being wrongfully dismissed as a mental health condition, leading to further stigmatization and inadequate treatment^33^. Finally, research into the underlying pathophysiology of PEM and development of effective treatments are needed to address not only the current burden of illness from COVID-19, but other known and emerging pathogens as well.

### 4.1. Strengths and limitations

This study was based on a large and diverse sample recruited primarily through address-based sampling and weighted to provide results generalizable to the NYC adult population, minimizing threats to external validity from selection and non-response bias. To our knowledge, this is the largest population-representative survey to screen for PEM symptoms. Nevertheless, several limitations should be considered when interpreting these results. First, the lack of a COVID-negative comparison group precludes direct attribution of PEM symptoms to SARS-CoV-2 infection. Second, participants with self-reported prior COVID-19 were included regardless of clinical or laboratory evidence. While this is appropriate for a population which experienced high transmission when testing was inaccessible, false positives or negatives may bias the results away from or towards the null, respectively. Third, while the DSQ-PEM has been shown to discriminate between infection-associated chronic illness patients and healthy controls in clinical samples, more evidence is needed to establish the specificity of the instrument in the general population to minimize false positives from people experiencing general fatigue rather than neuroimmune pathology. Fourth, while validated instruments were used for the primary measures in this study, the use of abbreviated versions may result in misclassification. In particular, the DSQ-PEM specifically screens for fatigue-related symptoms but may fail to capture more diverse PEM symptoms^34^. Finally, these results should not be interpreted as an estimate of clinically diagnosable PEM or ME/CFS. The DSQ-PEM alone is not a diagnostic instrument in the absence of clinical evaluation, and people who screen positive on the DSQ-PEM may not meet diagnostic thresholds on cardiopulmonary exercise testing^14,35^. Similarly, while symptom duration ≥ 14 hours is a diagnostic criterion for ME/CFS, our study did not assess other criteria such as orthostatic intolerance or unrefreshing sleep.

### 4.2. Conclusion

In conclusion, PEM symptoms were highly prevalent among adult NYC residents with a history of COVID-19, representing a significant and ongoing public health concern with particular implications for the long-term well-being of marginalized groups. To increase access to clinical care pathways and proper symptom management while reducing misdiagnosis and stigma, there is an urgent need for increased education and awareness of PEM among healthcare providers, health systems, public health agencies, and the communities they serve.

Author Contributions: Author contributions to this manuscript according to CRediT (Contributor Roles Taxonomy) roles are as follows: SEP – Conceptualization, Formal Analysis, Writing – original draft, Writing – review and editing; TR – Conceptualization, Writing – review and editing; JP – Validation, Writing – review and editing; JS – Conceptualization, Writing – review and editing; AL - Conceptualization, Writing – original draft, Writing – review and editing Disclosure of Funding and Conflicts of Interest: This work was supported by a Strengthening US Public Health Infrastructure, Workforce, and Data Systems grant from the Centers for Disease Control and Prevention (#NE11OE000057). The authors have no conflicts of interest to disclose.

## Data Availability

Data are restricted under data governance guidance and not made available to the public

