## Supplemental Tables for "Prevalence and Functional Outcomes of Post-Exertional Malaise among Adults with prior COVID-19: Results from a Representative Survey of New York City Residents"

Supplemental Information

### Table S-1a: Self-reported Laboratory and Clinical Evidence of Prior COVID-19 Infection From Study Eligibility Screener

|  | Frequency | Percent | Cumulative % |
| --- | --- | --- | --- |
| Positive PCR Test | 4,126 | 42.9 | 42.9 |
| Positive Rapid Test (No Positive PCR Test) | 3,844 | 40.0 | 82.8 |
| Positive Antibody Test (No Positive PCR or Rapid Test) | 398 | 4.1 | 87.0 |
| Healthcare Provider Suspected COVID-19 (No Positive Test) | 286 | 3.0 | 90.0 |
| Some Other Reason (No Clinical or Laboratory Evidence) | 966 | 10.0 | 100.0 |
| Note: Summary categories shown, as respondents could mark multiple options on eligibility screener.  Response options to the question “Do you think you have ever had COVID-19? (Select all that apply)” were:  “Yes, confirmed by a positive rapid test OR home test”; “Yes, confirmed by a positive PCR test at a testing site, clinic, or with a health care provider (usually takes 24-48 hours to get results)”; “Yes, a health care provider suspected I had COVID-19, but I did not take a test”; “Yes, based on an antibody test; Yes, for some other reason”; “No” | | | |

### Table S-2: Unadjusted and covariate-adjusted mean WHODAS-2 Scores by level of PEM Symptoms

|  | No PEM Symptoms | Mild/Moderate Duration | Prolonged Duration |
| --- | --- | --- | --- |
|  | Mean Score (95% CI) | Mean Score (95% CI) | Mean Score (95% CI) |
| a) Unadjusted Mean Scores |  |  |  |
| WHODAS-2 Global Score | 3.15 (2.95, 3.36) | 14.20 (13.42, 14.99) | 19.11 (17.27, 20.96) |
| Domain 1: Cognition | 0.48 (0.44, 0.52) | 2.38 (2.22, 2.54) | 3.05 (2.69, 3.41) |
| Domain 2: Mobility | 0.68 (0.63, 0.73) | 2.91 (2.73, 3.10) | 3.86 (3.47, 4.24) |
| Domain 3: Self-Care | 0.23 (0.20, 0.26) | 1.25 (1.11, 1.39) | 2.02 (1.60, 2.44) |
| Domain 4: Getting Along | 0.53 (0.49, 0.58) | 2.16 (1.98, 2.33) | 2.88 (2.47, 3.30) |
| Domain 5: Daily Life | 0.62 (0.57, 0.66) | 2.84 (2.69, 3.00) | 3.77 (3.39, 4.15) |
| Domain 6: Social Participation | 0.62 (0.58, 0.67) | 2.75 (2.57, 2.93) | 3.59 (3.23, 3.95) |
| b) Model-adjusted Mean Scores |  |  |  |
| WHODAS-2 Global Score | 3.64 (3.43, 3.85) | 12.45 (11.70, 13.20) | 17.17 (15.38, 18.97) |
| Domain 1: Cognition | 0.55 (0.51, 0.60) | 2.10 (1.95, 2.25) | 2.75 (2.40, 3.09) |
| Domain 2: Mobility | 0.80 (0.75, 0.85) | 2.55 (2.38, 2.72) | 3.45 (3.06, 3.83) |
| Domain 3: Self-Care | 0.29 (0.26, 0.33) | 1.04 (0.91, 1.18) | 1.79 (1.39, 2.19) |
| Domain 4: Getting Along | 0.60 (0.55, 0.65) | 1.90 (1.73, 2.07) | 2.60 (2.19, 3.00) |
| Domain 5: Daily Life | 0.71 (0.66, 0.75) | 2.52 (2.37, 2.67) | 3.39 (3.02, 3.76) |
| Domain 6: Social Participation | 0.71 (0.66, 0.76) | 2.43 (2.26, 2.60) | 3.23 (2.88, 3.59) |
| WHODAS-2 scores by PEM Symptoms. Adjusted mean scores are the predicted value derived from linear regression models adjusted for age group, gender, race/ethnicity, educational attainment, neighborhood poverty, number of COVID-19 infections, year of first infection, COVID-19 vaccination history, pre-existing chronic conditions, and pre-existing disability. | | | |

### Table S-3: Association of PEM Symptom Duration with Depression and Anxiety among participants with PEM symptoms

|  | | Probable Depression | | Probable Anxiety | |
| --- | --- | --- | --- | --- | --- |
|  |  | aRR (95% CI) | p | aRR (95% CI) | p |
| Unadjusted | PEM (Mild/Moderate Duration) | ref |  | ref |  |
|  | PEM (Prolonged duration) | 1.20 (0.99 , 1.46) | .067 | 1.27 (1.06 , 1.52) | .009 |
| Adjusted | PEM (Mild/Moderate Duration) | ref |  | ref |  |
|  | PEM (Prolonged duration) | 1.18 (0.98 , 1.43) | . 078 | 1.27 (1.07 , 1.51) | .006 |
| Models adjusted for age group, gender, race/ethnicity, educational attainment, neighborhood poverty, number of COVID-19 infections, year of first infection, COVID-19 vaccination history, pre-existing chronic conditions, and pre-existing disability. | | | | | |

### Table S-4: Association of PEM Symptom Duration with Activity Limitation among participants with PEM symptoms

| Domain of Activity Limitation | aMR | conf.low | conf.high | p.value |
| --- | --- | --- | --- | --- |
| WHODAS-2 Global Score | 1.33 | 1.18 | 1.47 | < 0.001 |
| Domain 1: Cognition | 1.30 | 1.13 | 1.47 | < 0.001 |
| Domain 2: Mobility | 1.29 | 1.15 | 1.44 | < 0.001 |
| Domain 3: Self-Care | 1.58 | 1.22 | 1.95 | < 0.001 |
| Domain 4: Getting Along | 1.31 | 1.10 | 1.52 | < 0.001 |
| Domain 5: Daily Life | 1.30 | 1.15 | 1.44 | < 0.001 |
| Domain 6: Social Participation | 1.29 | 1.14 | 1.44 | < 0.001 |
| Adjusted Mean Ratios (aMR) of WHODAS-2 scores for Prolonged Duration compared to Mild/Moderate Duration.  Models adjusted for age group, gender, race/ethnicity, educational attainment, neighborhood poverty, number of COVID-19 infections, year of first infection, COVID-19 vaccination history, pre-existing chronic conditions, and pre-existing disability. | | | | |

### Table S-5: Association of PEM Symptoms with Depression and Anxiety Among Participants with No History of Mood Disorders

|  |  | Probable Depression | Probable Anxiety |
| --- | --- | --- | --- |
|  |  | aRR (95% CI) | aRR (95% CI) |
| A | No PEM | ref | ref |
|  | PEM (Mild/Moderate Duration) | 5.10 (3.76 , 6.91) | 5.09 (3.89 , 6.67) |
|  | PEM (Prolonged duration) | 5.82 (3.74 , 9.06) | 7.19 (5.06 , 10.22) |
| B | No PEM | ref | ref |
|  | PEM (Mild/Moderate Duration) | 3.95 (2.89 , 5.40) | 3.91 (2.95 , 5.19) |
|  | PEM (Prolonged duration) | 5.08 (3.14 , 8.23) | 6.03 (4.08 , 8.91) |
| Notes:  Probable depression was defined as a PHQ-2 score ≥ 3. Probable Anxiety was defined as a GAD-2 score ≥ 3. All models were fit using modified Poisson regression for binary outcome data with robust standard errors and weighted to the target population of adult NYC residents with prior confirmed or suspected COVID-19.  Row A displays results from unadjusted models, while Row B displays results from models adjusted for age group, gender, race/ethnicity, educational attainment, neighborhood poverty, number of COVID-19 infections, year of first infection, COVID-19 vaccination history, pre-existing chronic conditions, and pre-existing disability. | | | |

### Figure S-1: Association of PEM Symptoms with Activity Limitations Among Participants with No History of Pre-Existing Disability

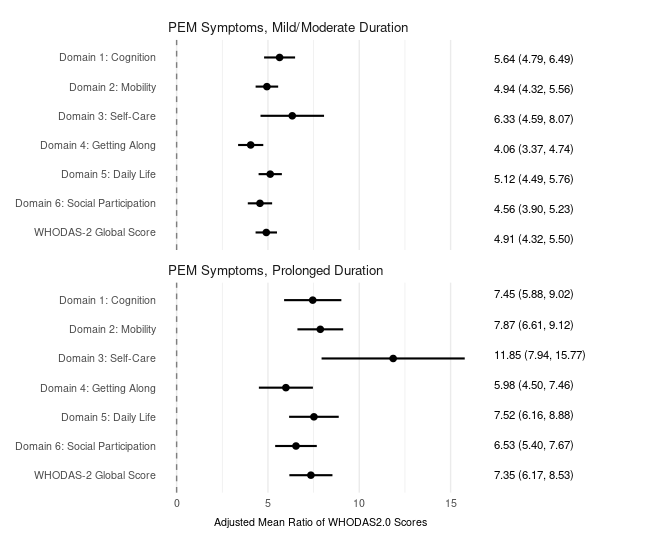
